# Patients’ perception of anaesthesia and participation in their own perioperative safety- An observational study

**DOI:** 10.1101/2025.02.05.25321349

**Authors:** Jyotsna Agarwal, Meena Nathan Cherian, Sana Yasmin Hussain, Pratibha Panjiar, Santosh Kumar, Davy Cheng

**Author notes:** Corresponding author: Davy Cheng (DC).

## Abstract

Patients for Patients Safety (PFPS) is a World Health Organization (WHO) program which advocates meaningful engagement of patients and families to improve patient safety. Perioperative patient safety encompasses the surgical risks, as well as risks due to anesthesia. This study was conceptualized to assess patient perception of their involvement in enhancing their own perioperative safety and also assess the current state of patient knowledge regarding anesthesia.

This observational cross-sectional study was conducted in a tertiary care teaching hospital of India. One hundred sixty-three consenting adults, aged 19 years to 75 years, scheduled for elective surgery, between May and July 2022, having American Society of Anesthesiologists grade 1 to 3 participated in the study. Patients having psychiatric disorder, cognitive impairment, speech and hearing problems were excluded.

A pre-validated questionnaire of 21 questions was administered in pre-operative area. Data collection was done in online MS excel spreadsheet format.

In our study it was encouraging to note that the general awareness on requirement of anesthesia for surgery was widely acknowledged. Nonetheless, the information about anesthesia as a separate specialty, and anesthesiologists as specialized doctors, was lacking (43/163, 28.8%). The inadequate awareness about anesthesia specialty as found in our study, might not be a true reflection of a wider population, which calls attention to invest in anesthesia awareness programs. Our study revealed a very promising initial step in patient empowerment, as majority of patients (100/163; 61.3%) were amenable to participate in their own perioperative safety. Regardless of education status, the population in general needs to be familiarized with the shared patient responsibility in enhancing patient safety, just like other public health programs. Our study aligns with the recently proposed concrete interventions for improving quality of perioperative care at a global stage which contributes to Sustainable Development Goals targets towards achieving Universal Health Coverage.

## Introduction

Patients for Patients Safety (PFPS) is a World Health Organization (WHO) program which advocates meaningful engagement of patients and families to improve patient safety. Patient engagement is described as “What can patients do to prevent medical mistakes?” a prominent initiative which was developed as a part of the patient safety movement [1]. Patient engagement has been shown to be vital to achieve integrated and people-centered health systems and services [2,3].

PFPS concept was first introduced in 2005 [2]. Even almost two decades later, the awareness of patients with regards to the implication of PFPS in perioperative safety, is unexplored. It would be pragmatic to understand the patient’s perception of perioperative safety and their understanding of the shared responsibility of patients in enhancing their own perioperative safety. This knowledge would further assist us in designing awareness programs for the community. Patients’ comprehension of their role in improving perioperative safety would eventually translate into patient cooperation, mutual collaboration amongst patients and medical staff, and furthermore, can reduce incidences of violence against healthcare delivery systems.

Perioperative patient safety encompasses patient safety from the surgical risks, as well as from the risks due to anaesthesia. Recent study revealed postoperative deaths accounts for 7.7% of all deaths globally, making it the third greatest contributor to deaths, after ischaemic heart disease and stroke [4]. A landmark systematic review and meta-analysis of 21.4M general anesthesia for surgery, found that mortality solely attributable to anesthesia has significantly declined over time, from 357 per million before the 1970s to 34 per million in the 1990s-2000s. Total perioperative mortality also decreased, with the greatest decline observed in developed countries; however, perioperative mortality rates were higher in developing countries compared to developed ones, indicating a disparity in healthcare quality and access [5]. Studies over the years, have revealed variable degree of awareness about anaesthesia amongst the patients, families and community [6–8]. Post-COVID epidemic, awareness about anaesthesia specialty and the role of anaesthesiologists in intensive care units, has improved. However, still, there is a wide gap in the knowledge about anaesthesia and perioperative safety in Low-and Middle-Income Countries [9].

On the backdrop of this information and beholding the vision of PFPS, this study was conceptualized, to assess patient perception of their involvement in enhancing their own perioperative safety. Additionally, the current state of patient knowledge and apprehensions regarding anaesthesia was assessed.

## Methodology

Ethics approval was given by Jamia Hamdard Institutional Ethics Committee (JHIEC 01/20) and the study was registered prospectively with Clinical Trials Registry-India (CTRI/2020/10/028318, http://ctri.nic.in).

This observational, cross-sectional study was conducted over a period of three months, in a tertiary care teaching hospital in North India. All participants gave written informed consent before taking part in the study. The pilot testing of the questionnaire was done in October 2020. However, due to COVID the study was discontinued temporarily, the study resumed on 2nd May 2022 and completed on 30th July 2022. One hundred sixty-three consenting adults, aged 19 years to 75 years, scheduled for elective surgery, having American Society of Anesthesiologists grade 1 to 3 participated in the study. Patients having psychiatric disorders, cognitive impairment, speech and hearing problems were excluded.

Questionnaire was prepared by all the study investigators. The questionnaire was assessed and validated by various faculties which included anesthesiologists, surgeons and psychiatrists. The questionnaire assessors were not a part of the study. Each study question was assessed under the following headers: relevance, clarity, simplicity and non-ambiguity.

The questionnaire was in English (available as supplementary material), which was administered to the patients by a dedicated interviewer in the preoperative area. The interviewer was an anesthesiologist. The questions were asked in English or in the local language Hindi, as per patients’ understanding. The interviewer was not involved in any subsequent steps of the study. The questionnaire also contained some open-ended questions to allow flexibility for the participants to add their own insights. After completing the questionnaire, the interviewer addressed patients’ doubts and educated the patients on shared responsibility in their own perioperative safety.

The questionnaire consisted of 21 questions divided into 4 sections and took ten minutes to complete. Section 1 contained questions related to patient demography, education status and prior exposure to anesthesia and surgery. Section 2 explored patients’ awareness about anesthesia, anesthesiologists, pre-anesthesia examination, patients’ understanding of their relevant health conditions and relevance of ‘nil per oral’ status before surgery. Section 3 assessed patients’ concerns such as fear of unconsciousness during surgery, pain in perioperative period and any other concerns such as anxiousness and confidence regarding the surgery. Section 4 focused on patients’ perception of their participation in enhancing their own perioperative safety (PFPS).

The collected data were pooled and coded in MS Excel spreadsheet program. SPSS v23 (IBM Corp.) was used for data analysis. Descriptive statistics were presented as means ± standard deviations, medians ± IQRs and in graphical manner using histograms/box-and-whisker plots/column charts for continuous variables. Data was presented as frequencies and percentages and graphically as bar charts/pie charts for categorical variables. Group comparisons for continuously distributed data were made using independent sample ‘t’ test when comparing two groups. Appropriate non-parametric tests in the form of Wilcoxon Test were used for non-normally distributed data. Chi-squared test was used for group comparisons for categorical data. In case the expected frequency in the contingency tables was found to be <5 for >25% of the cells, Fisher’s Exact test was used instead. Linear correlation between two continuous variables was explored using Pearson’s correlation (if the data were normally distributed) and Spearman’s correlation (for non-normally distributed data). Statistical significance was kept at p < 0.05. The AUROC were compared using DeLong’s method.

## Results

A total of 163 patients participated. Mean age of patients was 36.61 ± 13.11 years, 61 (37.4%) patients were males and rest were females. Almost half of the patients had attended at least primary school (78 /163;47.9%), one fourth did not receive even primary school education (42/163; 25.8%), whereas remaining one fourth received education beyond high school (43/163;26.4%) (Table 1).

**Table 1.** Summary of all parameters.

| Basic Details | Mean ± SD \|\| Median (IQR) \|\| Min-Max \|\| Frequency (%) |
| --- | --- |
| **Age (Years)** | 36.61 ± 13.11 \|\| 35.00 (27.00-46.00) \|\| 19.00 - 75.00 |
| **Age** |  |
| 19-30 Years | 71 (43.6%) |
| 31-40 Years | 36 (22.1%) |
| 41-50 Years | 28 (17.2%) |
| 51-60 Years | 18 (11.0%) |
| 61-70 Years | 9 (5.5%) |
| 71-80 Years | 1 (0.6%) |
| **Gender** |  |
| Male | 61 (37.4%) |
| Female | 102 (62.6%) |
| **Educational Level** |  |
| No schooling | 42 (25.8%) |
| Received Primary School Education | 78 (47.9%) |
| Received Education Beyond School | 43 (26.4%) |
| **Had experience of Previous Surgery** | 59 (36.2%) |
| **Had experience of Previous Anaesthesia** |  |
| Needle in Back | 30 (50.8%) |
| Full Unconsciousness | 25 (42.4%) |
| Local Anaesthesia | 2 (3.4%) |
| Full Unconsciousness, Needle in back | 1 (1.7%) |
| Don't know | 1 (1.7%) |
| **Who Gives Anesthesia** |  |
| Don't know | 86 (52.8%) |
| Specialist Doctor | 47 (28.8%) |
| Surgeon | 20 (12.3%) |
| Nurse | 7 (4.3%) |
| Technician | 3 (1.8%) |
| **Confidence Regarding Perioperative Safety** |  |
| Fully Confident | 70 (42.9%) |
| Not Confident | 93 (57.1%) |
| **Reason for 'Nil per Oral'** |  |

| Basic Details | Mean ± SD \|\| Median (IQR) \|\| Min-Max \|\| Frequency (%) |
| --- | --- |
| Don't know | 115 (70.6%) |
| Helps in surgery | 42 (25.8%) |
| Helps in safe Anesthesia and prevent airway complications | 6 (3.6%) |
| **Know If Anaesthesia Is Required for the Planned Surgery** |  |
| Yes | 132 (81.0%) |
| Don't Know | 31 (19.0%) |
| **Knows the Word Anaesthesia** |  |
| Yes | 39 (23.9%) |
| No | 124 (76.1%) |
| **Can Help Improving Own Perioperative Safety** |  |
| Yes | 100 (61.3%) |
| No | 40 (24.5%) |
| Don't Know | 23 (14.1%) |
| **Know About Things to Inform Before Surgery** |  |
| Yes | 104 (63.8%) |
| Don't Know | 59 (36.2%) |
| **Know Whether PAE was done** |  |
| Yes | 114 (69.9%) |
| Don't Know | 49 (30.1%) |
| **Wanted to Know About After-Care** |  |
| Yes | 85 (52.1%) |
| No | 78 (47.8%) |

### Awareness regarding anaesthesia

Although majority of patients (132/163; 80.9%) knew that some kind of unconsciousness or numbness (anaesthesia) will be provided to them during surgery, only one third of the study participants (43/163, 28.8%) acknowledged that anaesthesia is administered by a specialist doctor. Others guessed that it could be administered by the surgeon, nurse or technician. All patients knew the word ‘surgery’, only 24.0% patients knew the word ‘anaesthesia’ (Table 1).

Pre-anaesthesia evaluation (PAE) was done for all patients who were surveyed in pre-operative area. However, only 70% (114/163) patients were aware that their PAE was done (Table 1). One fourth (42 /163) patients thought that ‘nil per oral’ (NPO) is a requirement for surgery, and less than 4% patients were aware that NPO status is also a requirement for safe anaesthesia (Table 1). No significant association was found between awareness about NPO status with educational level, awareness about anaesthesia, previous surgery, prior PAE evaluation.

Sixty-four percent (104 / 163) patients said that they should inform relevant things to the doctor. Out of the 104 patients, majority of patients said that informing to the doctor of their current co-morbidities (83.6%; 87/ 104, p<0.001) and previous surgeries (43.26%; 45 /104, p<0.001) was relevant for their perioperative safety. Patients also addressed that informing about their current medications and any allergies, including drug allergies is relevant (Table 2, Table 3).

**Table 2.** Distribution of all parameters.

| **Comorbidities present** | **Yes** | **No** |
| --- | --- | --- |
| **Health Condition** | 44 (27.0%) | 119 (73.0%) |
| **Medication** | 23 (14.1%) | 140 (85.9%) |
| **Allergy** | 10 (6.1%) | 153 (93.9%) |
| **Pregnancy** | 1 (0.6%) | 162 (99.4%) |
| **Smoking** | 11 (6.7%) | 152 (93.3%) |
| **None** | 104 (63.8%) | 59 (36.2%) |
| **Others** | 0 (0.0%) | 163 (100.0%) |
| **Concerns in perioperative period** | **Yes** | **No** |
| **No concerns / Fully confident** | 70 (42.9%) | 93 (57.1%) |
| **Unconsciousness during surgery** | 24 (14.7%) | 139 (85.3%) |
| **Wake up Time after surgery** | 27 (16.6%) | 136 (83.4%) |
| **Comorbidities present** | **Yes** | **No** |
| Pain | 29 (17.8%) | 134 (82.2%) |
| Anxiety, unknown fear | 23 (14.1%) | 140 (85.9%) |
| Successful outcome of surgery | 40 (24.5%) | 123 (75.5%) |
| **Can Help Improving Own Safety** | **Yes** | **No** |
| Yes, By not Being Anxious | 48 (29.4%) | 115 (70.6%) |
| Yes, By taking Care of Health | 5 (3.1%) | 158 (96.9%) |
| Yes, By co-operating with doctor | 52 (31.9%) | 111 (68.1%) |
| Yes, by telling relevant Things | 7 (4.3%) | 156 (95.7%) |
| Yes, but Don't Know How | 7 (4.3%) | 156 (95.7%) |
| No, because God Is Everything | 8 (5.0%) | 155 (95.0%) |
| No, because Doctor Is responsible | 32 (19.6%) | 131 (80.4%) |
| **Things to Inform to the doctor before surgery** | **Yes** | **No** |
| Correct Site | 1 (0.6%) | 162 (99.4%) |
| Health Condition (co-morbidities) | 87 (53.4%) | 76 (46.6%) |
| Medications | 30 (18.4%) | 133 (81.6%) |
| Allergy | 30 (18.4%) | 133 (81.6%) |
| Pregnancy | 3 (1.8%) | 160 (98.2%) |
| Smoking | 7 (4.3%) | 156 (95.7%) |
| Previous Surgery | 45 (27.6%) | 118 (72.4%) |
| **Concerns regarding care post-surgery (after care)** | **Yes** | **No** |
| Diet | 24 (14.7%) | 139 (85.3%) |
| Stitches | 11 (6.7%) | 152 (93.3%) |
| Other Precautions | 13 (8.0%) | 150 (92.0%) |
| Medications | 8 (4.9%) | 155 (95.1%) |
| Normal Activities | 39 (23.9%) | 124 (76.1%) |
| Child Care | 5 (3.1%) | 158 (96.9%) |
| Discharge | 15 (9.2%) | 148 (90.8%) |
| Complications | 16 (9.8%) | 147 (90.2%) |

**Table 3:** Association between ‘Things to Inform Before Surgery and Parameters’.

|  |  |  |
| --- | --- | --- |
| **Parameters** | **Know About Things to Inform Before Surgery** | **p value** |

|  | Yes<br>(n = 104) | Don't Know<br>(n = 59) |  |
| --- | --- | --- | --- |
| Things to Inform: Correct Site (Yes) | 1 (1.0%) | 0 (0.0%) | 1.000<sup>2</sup> |
| Things to Inform: Health Condition (Yes)*** | 87 (83.7%) | 0 (0.0%) | <0.001<sup>1</sup> |
| Things to Inform: Medications (Yes)*** | 30 (28.8%) | 0 (0.0%) | <0.001<sup>1</sup> |
| Things to Inform: Allergy (Yes)*** | 30 (28.8%) | 0 (0.0%) | <0.001<sup>1</sup> |
| Things to Inform: Pregnancy (Yes) | 3 (2.9%) | 0 (0.0%) | 0.554<sup>2</sup> |
| Things to Inform: Smoking (Yes)*** | 7 (6.7%) | 0 (0.0%) | 0.049<sup>2</sup> |
| Things to Inform: Previous Surgery (Yes)*** | 45 (43.3%) | 0 (0.0%) | <0.001<sup>1</sup> |

\*\*\*Significant at $p < 0.05$ , 1: Chi-Squared Test, 2: Fisher's Exact Test

### Concerns regarding perioperative period

One third patients (51/163; 31.2%) were concerned regarding unconsciousness during surgery and waking up after surgery. Pain after surgery was worrisome for 18% patients (Table 2). In aftercare, most patients wanted to know about diet (14.7%) and resumption of normal activities (23.9%) after surgery (Table 2). Almost half of the surveyed patients (70/163; 42.9%) were confident regarding their surgery and had no perioperative concerns (Table 1).

### Patient participation in their own safety (Patients for Patient Safety)

A very positive finding of the study was that more than half (100/163; 61.3%) of the surveyed patients agreed that they can help in improving their own perioperative safety (Table 1).

## Discussion

Total 163 patients were surveyed on their perception about participation in their own perioperative safety. Additionally, awareness about anesthesia specialty and their apprehensions regarding anesthesia were also revealed. For patient engagement in their own perioperative safety, information regarding anesthesia is as important as is information regarding surgery.

### Patient participation in their own perioperative safety

More than half of the surveyed patients were cognizant of their participation in improving their own perioperative safety (Table 1). This, they believed, can be done by cooperating with the doctors, following their instructions, taking prescribed medications and staying calm (Table 2). Also, these patients were significantly more aware about anaesthesia and relevant history which should be told to their doctors before surgery such as allergies, co-morbidities and previous surgical exposure (Table 4). However, this did not correlate with educational status or prior exposure to surgery and anaesthesia.

**Table 4:** Association between ‘Participation in Own Safety and Parameters’.

| Parameters | Can Help Improving Own Safety |  |  | p value |
| --- | --- | --- | --- | --- |
|  | Yes<br>(n = 100) | No<br>(n = 40) | Don't Know<br>(n = 23) |  |
| **Know If Anaesthesia Is Required\*** |  |  |  | <0.001<sup>1</sup> |
| Yes | 90 (90.0%) | 26 (65.0%) | 16 (69.6%) |  |
| Don't Know | 10 (10.0%) | 14 (35.0%) | 7 (30.4%) |  |
| **Knows the Word Anaesthesia / Anaesthetist (Yes) \*** | 36 (36.0%) | 2 (5.0%) | 1 (4.3%) | <0.001<sup>1</sup> |
| **Know About Things to Inform Before Surgery\*** |  |  |  | <0.001<sup>1</sup> |
| Yes | 77 (77.0%) | 14 (35.0%) | 13 (56.5%) |  |
| Don't Know | 23 (23.0%) | 26 (65.0%) | 10 (43.5%) |  |
| **Things to Inform: Health Condition (Yes) \*** | 66 (66.0%) | 10 (25.0%) | 11 (47.8%) | <0.001<sup>1</sup> |
| **Things to Inform: Allergy (Yes) \*** | 25 (25.0%) | 2 (5.0%) | 3 (13.0%) | 0.017<sup>1</sup> |
| **Things to Inform: Previous Surgery (Yes) \*** | 36 (36.0%) | 3 (7.5%) | 6 (26.1%) | 0.003<sup>1</sup> |

\*Significant at $p < 0.05$ , 1: Chi-Squared Test

Notably, one fourth (40/163, 24.5%) of the surveyed patients said that they had no role in their own perioperative safety (Table 1). Out of these 40 patients, eight patients believed that perioperative outcomes are entirely in the ‘hands of God’ whereas 32 patients (32/163, 19.6%) felt that perioperative outcome lies entirely in the ‘hands of the doctors’ (Table 2). Notably, 20% that is one-fifth of the surveyed patients believed that their safety lies entirely in the ‘hands of doctors’. This cohort can potentially turn hostile to the medical team in the event of unfavorable perioperative outcomes, which could be misrepresented through social media. Therefore, it is important to talk with patients’ community and on social media platforms about PFPS awareness. Pre-operative period does offer a ‘teachable’ moment when these patients and their families can be identified and educated on shared patient responsibility in perioperative safety.

In our study, we found that patients in spite having had prior exposure to anesthesia and surgery, patients were not aware of the concept of participation in their own perioperative safety. This may reflect the lack of effective communication by the perioperative team. Therefore, effective communication by the perioperative team can have a significant impact as revealed by a study by Wolter CB et al [10] on patients’ perspectives on patient engagement in their safety. They found that during communication regarding patient safety, patients related more to terms like ‘your safety’ instead of ‘patient safety’ and thought engagement in healthcare decisions is their ‘right’. However, they believed the ultimate responsibility of their safety, should still lie, in the hands of healthcare professionals [10]. This finding resonates with our study population who were of the opinion that patient safety lies entirely in the ‘hands of the doctors’.

Our study is in alignment with the findings of Gobbo et al11, which noted that patients value pre-admission contact, specific education, effective communication, continuity of care, and privacy during the perioperative period. Our study agrees that these are crucial factors for enhancing patient satisfaction and safety [11].

Like our study, Dixon and Tillman [12] did not find correlation of education level with the patients’ perception of their safety. Neither did modern patient safety initiatives directly influence the patients’ perception. Physician trust and patient communication was found to have the greatest impact on the patients feeling safe, before surgery. It has been documented that patients who had more trust in the doctor and had better communication from the health care workers were 30% less likely to raise a complaint against the doctor [12,13].

### Awareness about anaesthesia

We found that more than 80% of the surveyed patients were aware that they would receive some kind of numbness or unconsciousness before their surgery, however, the majority were oblivious about anesthesia being administered by specialized doctors (Table 1). Similar findings have been reported in previous studies. Despite a high level of awareness (82-92%) regarding the need of some kind of anesthesia for surgery,[8,14,15] the knowledge about anesthesia being a separate specialty and anesthesiologists being specialized doctors was found to be limited (30-50%) [6,8,14].

Contrary to our study, some studies have found high levels of awareness about anesthesiologists. A survey conducted in visitors to a health fair in Bangalore, India in 2014,15 revealed very high awareness (75% of participants) about anesthesiologists being doctors. This could be because the survey was conducted in a specific cohort of educated industry workers, and the questionnaire design had leading questions with binary response format. Similarly high level of awareness amongst patients about anesthesiologists as ‘specialized doctors’ was observed in a study from Ghana [16]. This was attributed to the fact that 88% surveyed patients were educated, and they were referred to a ‘special doctor’ (anesthetist) by the surgeon. This highlights the role of team communication, including the surgical colleagues, in creating anesthesia awareness.

In our study, only 24% of patients knew the word ‘anesthesia’ (Table 1), however, most of the patients could relate to the more commonly used term ‘behoshi’ which in the local language which means ‘unconsciousness’. Similarly, unawareness of the term ‘anesthesiologist’ was observed as compared to ‘physicians’ and ‘surgeons’ by Prasad et al [15]. We feel that this lack of awareness of specific terminology for unconsciousness might have impacted awareness about anesthesia as an independent specialty and anesthesiologists as independent doctors, at par with surgery and surgeons. Surprisingly, in a study conducted in 2009, awareness about spectrum of anesthesia procedures available to the patient was found lacking not only in general population, but also amongst educated people including medical undergraduate students [17].

Also, in our study, educated patients were significantly more aware about the term anesthesia, need of anesthesia for the surgery, and were more engaged in their after-care requirements, as compared to uneducated patients (Table 5). A positive correlation of education and anesthesia awareness is well-known [6,8,14,18,19].

**Table 5:** Association between ‘Educational Level’ and Parameters.

| Parameters | Educational Level |  |  | p value |
| --- | --- | --- | --- | --- |
|  | No schooling<br>(n = 42) | Received School<br>Education<br>(n = 78) | Received Education<br>Beyond School<br>(n = 43) |  |
| **Know If Anaesthesia Is Required\*** |  |  |  | 0.003<sup>1</sup> |
| Yes | 28 (66.7%) | 63 (80.8%) | 41 (95.3%) |  |
| Don't Know | 14 (33.3%) | 15 (19.2%) | 2 (4.7%) |  |
| **Knows the Word Anaesthesia /**<br>**Anaesthetist (Yes) \*** | 0 (0.0%) | 14 (17.9%) | 25 (58.1%) | <0.001<sup>1</sup> |
| Yes | 24 (57.1%) | 47 (60.3%) | 33 (76.7%) |  |
| Don't Know | 18 (42.9%) | 31 (39.7%) | 10 (23.3%) |  |
| **Know Reason for Fasting (Yes) \*** | 6 (14.3%) | 27 (34.6%) | 15 (34.9%) | 0.044<sup>1</sup> |
| **Wanted to Know About After-Care (Yes) \*** | 12 (28.6%) | 46 (59.0%) | 27 (62.8%) | 0.002<sup>1</sup> |

\*Significant at $p < 0.05$ , 1: Chi-Squared Test

The lack of effective communication by the anesthesia team is reflected in the following findings of our study. Previous experience of surgery did not result in enhanced awareness about anesthesia. Although all patients had undergone Pre-Anesthesia Evaluation (PAE), still one third of them were not aware that they had completed PAE. Less than 4 % patients knew that recommended ‘nil per oral’ (NPO) status is vital for safe anesthesia delivery (Table 1). Many patients believed that preoperative fasting was a requirement for abdominal surgeries only. This lack of awareness could lead to non-compliance with preoperative NPO instructions in patients posted for non-abdominal surgeries, leading to adverse anesthesia outcomes.

Lack of correlation of anesthesia awareness with the history of previous surgery has been seen in earlier studies as well [6,14]. The necessity to improve patient communication by anesthesia team, particularly during preoperative visits, can be an effective measure [6,8,18,19].

Indian Society of Anaesthesiologists have designed an ‘ISA Public Awareness Flier’ [20]. It has two sections ‘Know your Anaesthesia’ and ‘Know your Anaesthesiologist’, which is a very useful tool for spreading anaesthesia awareness among patients and their family. This can be made available in PAE clinics and surgical out-patient clinics and wards.

### Things to inform before surgery

Sixty-four percent patients were aware of the importance of informing relevant things to the doctor preoperatively, particularly their current co-morbidities and previous surgeries (Table 1, Table 2). Notably only one patient of the entire sample size (1 out of 163; 0.6%) (Table 2) said that correct site of surgery must be emphasized. Although patients are aware that wrong site surgeries can happen erroneously, they also need to be educated about the role of patient participation in reducing such errors.

### Concerns regarding perioperative period

The main concerns of patients during perioperative period were regarding successful surgery, unconsciousness during surgery, waking up after surgery, pain management after surgery, resumption of diet and normal activities after surgery. A substantial proportion of patient population which comes to our hospital are daily wages workers, therefore resumption of a normal diet and activities also reflect returning to work early and earn. Notably, half of the patients (80/ 163; 49 %) were mainly concerned about anaesthesia related outcomes such as unconsciousness during surgery (24/163), waking up after surgery (27/163) and pain management after surgery (29/163) (Table 2).

In our study almost half (42.9%) of the patients were confident regarding their surgery and did not have any perioperative concerns or fears. An important factor for this patient confidence was the patient faith on their surgeon. As noted by Dixon and Tillman [12], physician trust and patient communication have a huge impact on patients feeling safe before surgery.

Previous studies have reported similar findings of fear of unconsciousness during surgery, not waking up from anaesthesia after surgery, pain during and after surgery, and fear of death [6,18]. Role of public health education programs and using media for education have been suggested [6,17,18]. Our study highlights the importance of integrated programs for improved communication in PAE which can alleviate such patient fears and encourage patient empowerment.

In our study, educated patients were found to be generally more aware of anaesthesia, they had more concerns regarding perioperative period and knew that they need to tell relevant things to the doctor before surgery (Table 5). Despite this enhanced awareness about anaesthesia and perioperative care, educated patients were unaware about patient participation in enhancing perioperative safety. As precisely noted by Wolter CB et al [10], although patients demand engagement in healthcare decisions but believe that the ultimate responsibility of their safety still lies in the hands of healthcare professionals only, which is reflected in our study also.

One limitation of our study is the small sample size; therefore, the outcome represents the perception of a small cohort of patients. Future studies in larger population are needed to explore the concept of patient engagement in perioperative safety.

In conclusion, it was encouraging to note in our study that the general awareness on requirement of anesthesia for surgery was widely acknowledged. Nonetheless, the perception about anesthesia as a separate specialty, and anesthesiologists as specialized doctors, was lacking. The inadequate awareness about anesthesia specialty as found in our study, might not be a true reflection of a wider population, which calls attention to invest in ‘anesthesia awareness programs’. This should be addressed globally through academia, professional societies, public health and social platforms. To the best of our knowledge, none of the previous studies have explored the patient perception of ‘Patients for Patient Safety’ in the perioperative period. Our study revealed a very promising initial step in patient empowerment, that majority of patients were amenable to participate in their own perioperative safety. Regardless of education status, the population in general needs to be familiarized with the shared patient responsibility in enhancing patient safety, just like other public health programs. High-quality surgical services that ensure patient safety are essential to achieve the health-related targets of the United Nations Sustainable Development Goals [21]. Our study also aligns with the proposed concrete interventions for improving quality of perioperative care at a global stage towards achieving Universal Health Coverage [22].

## Supporting information

Questionnaire

STROBEchecklist

## Data Availability

The data presented in this article are openly available and have been uploaded as supplementary materials. This study adheres to the STROBE guidelines and has been registered with a clinical trial registration number. [Institutional Ethics committee: JHIEC 01/20. Clinical Trials Registry-India: CTRI/2020/10/028318.]

## Contributorship statement

- Substantial contributions to the conception or design of the work; or the acquisition, analysis, or interpretation of data for the work; AND
- Drafting the work or revising it critically for important intellectual content; AND
- Final approval of the version to be published; AND
- Agreement to be accountable for all aspects of the work in ensuring that questions related to the accuracy or integrity of any part of the work are appropriately investigated and resolved.

All authors have contributed to the above 4 criteria of authorship.

Contribution of authors in the study

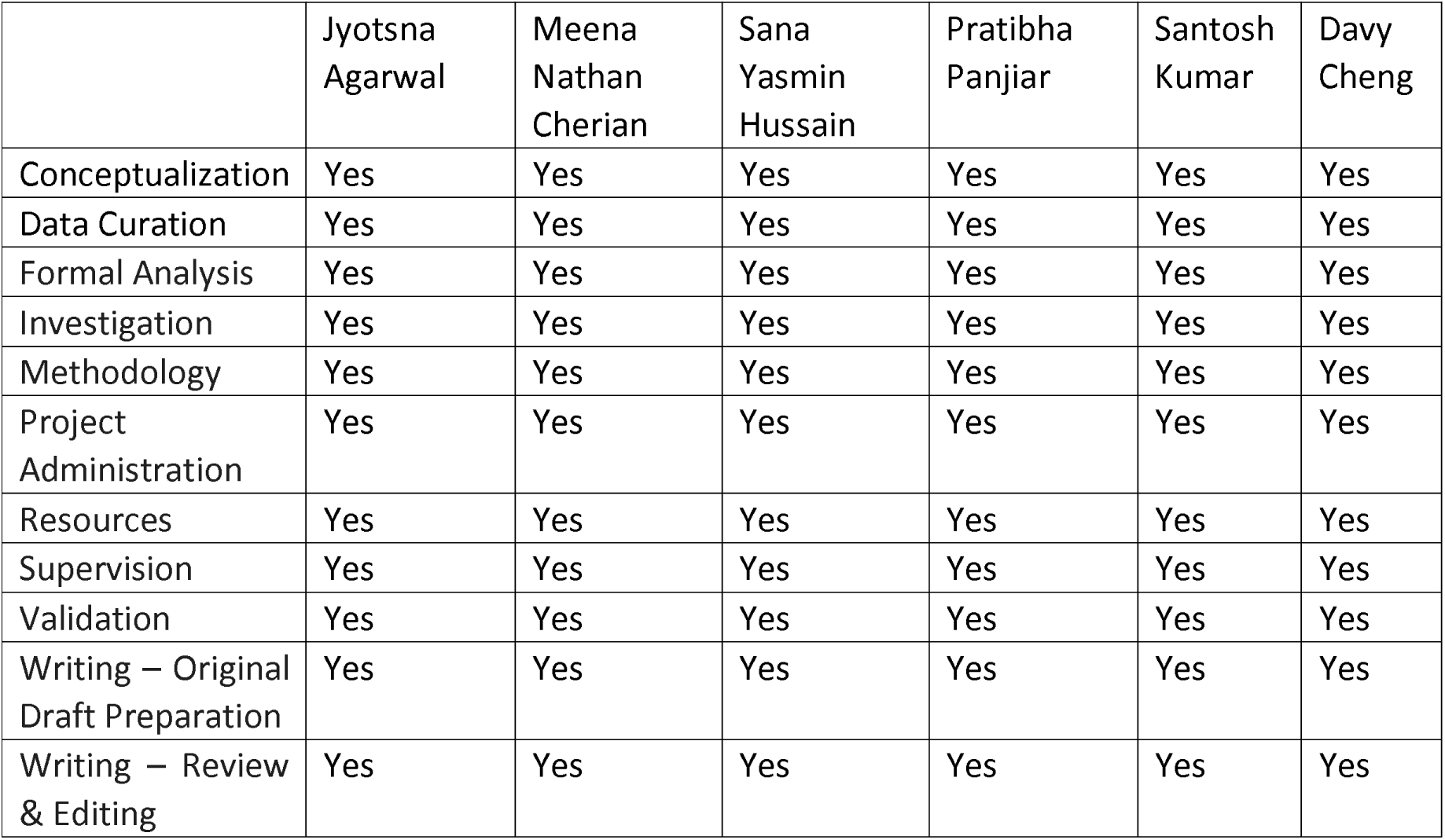

The authors declare that there is no conflict of interest and no external funding was received for the study.

