## Supplementary material for "Patients’ perception of anaesthesia and participation in their own perioperative safety- An observational study": Questionnaire

|  | Question | Options | Remarks |
| --- | --- | --- | --- |
| Section 1 | **Patient demography** | | |
| 1. | Date of filling the form |  |  |
| 2. | Hospital admission number |  |  |
| 3. | Name of the planned surgical procedure |  |  |
| 4. | Age |  | Only whole numbers to be written. |
| 5. | Gender | 1. Male 2. Female 3. Others |  |
| 6. | Educational level | 1. No schooling  2. Primary school education  3. Education beyond school |  |
| 7. | Does the patient has any of the following history  (Comorbidities) | 1. Health conditions ( suchas diabetes, high   blood pressure, thyroid diseases, breathing difficulty, and heart, kidney, liver, neurological, psychological illness)   2. Current medications including herbal medications 3. Allergies 4. Pregnancy and Breast feeding 5. Smoking, tobacco and alcohol intake, drug addiction 6. None of the above 7. Other | Tick all the applicable options. Any additional information to be recorded in ‘other’. |
| 8. | Have you had any previous surgery ? | 1. Yes  2. No |  |
|  | If answer to previous question is Yes, then, Did you receive anaesthesia for your **previous** surgery ? | 1.Don’t know  2. No  3. Full unconsciousness was given  4. Needle in back was inserted for anaesthesia  5. Any other comments ­ | Attempt if answer to previous question was ‘Yes”  Mark all options that the patient says. The patient might have had more than one previous surgeries. |
| Section 2 | **Patient awareness about anaesthesia and anaesthesiologist** | | |
| 9. | Have you heard the word 'Anaesthesia" | 1. Yes 2. No |  |
| 10. | Do you know if anaesthesia is required for your **present** surgery ? | 1. Yes 2. No 3. Don’t know |  |
| 11. | Do you know who gives anaesthesia ? | 1. Don’t know  2. Surgeon  3. Specialist doctor in  anaesthesia  4. Nurse  5. Technician  Any other comments | Tick all the options mentioned by the patient. Any additional information to be recorded in ‘any other comments’. |
| 12. | Were you examined by a doctor for ‘Pre Anesthesia Examination’(PAE) for this surgery ? | 1. Yes 2. No 3. Don’t know | All patients had undergone PAE for the planned surgery. Interviewer must confirm this from patient file. |
| 13. | Do you know that there are some relevant things to tell your doctor before surgery ? | 1. I don’t know 2. Yes, I know I should tell some relevant things about myself to the doctor |  |
| 14. | If answer to previous question is Yes, then  What are the things you should tell about you to your doctor before surgery? | I know that I should tell my doctor before surgery about my   1. Correct site of surgery 2. Health conditions (diabetes, high blood pressure, thyroid diseases, breathing difficulty, and heart, kidney, liver, neurological, psychological illness) 3. Current medications including herbal remedies 4. Allergies 5. Pregnancy and breast feeding 6. Smoking, tobacco and alcohol intake, drug addiction 7. Previous surgery and anaesthesia   Any other comments | Tick all the options mentioned by the patient. Any additional information to be recorded in ‘any other comments’. |
| 15. | Do you know why you should remain ‘nil per oral’ before this surgery ? | 1. Don’t know 2. Helps in surgery 3. Helps in safe anaesthesia, prevents airway complications   Any other comments | Tick all the options mentioned by the patient. Any additional information to be recorded in ‘any other comments’ |
| Section 3 | **Patient concerns regarding perioperative period** | | |
| 16. | How confident you are feeling regarding your safety in undergoing this surgery ? | 1. Not confident 2. Somewhat confident 3. Fully confident |  |
| 17. | Do you have any concerns about this surgery ? | 1. I have no concerns 2. Will I be conscious during surgery 3. When will I wake up after surgery 4. Will I have pain   Any other comments | Tick all the options mentioned by the patient. Any additional information to be recorded in ‘any other comments’. |
| 18. | Do you want to know anything regarding care after your surgery ? | 1. Yes, I want to know some things 2. No |  |
| 19. | If answer to previous question is Yes, then what do you want to know regarding care after the surgery | I want to know about   1. precautions in eating and drinking 2. when will the stitches be removed (wound care) 3. things to avoid after surgery 4. medications to be taken 5. resumption of normal activities (job etc) 6. child care , breast feeding, pregnancy 7. discharge 8. other complications | Tick all the options mentioned by the patient. Any additional information to be recorded in ‘other complications’. |
| Section 4 | **Patients for Patient Safety** | | |
| 20. | Do you think you can help in improving your own safety related to this surgery? | 1. Yes  2. No  3. Don’t know |  |
| 21. | If answer is Yes / No to previous question, then explain why / how ? | Open ended question | Write answers in patients words |
